# Ambulatory Duchenne Muscular Dystrophy Children: Cross-sectional Correlation between Function, Quantitative Muscle Ultrasound and MRI

**DOI:** 10.1101/2021.08.17.21262119

**Authors:** Hala Abdulhady, Hossam M. Sakr, Nermine S. Elsayed, Tamer A. El-Sobky, Nagia Fahmy, Amr M. Saadawy, Heba Elsedfy

## Abstract

**Introduction/Aims:** Duchenne muscular dystrophy (DMD) is a progressive genetic muscle disease. Quantitative muscle ultrasound (MUS), muscle MRI, and functional tools are important to delineate characteristics of muscle involvement. We aimed to establish correlations between clinical/functional and above-named imaging tools respecting their diagnostic and prognostic role in DMD children.

**Methods:** A Prognostic cross-sectional retrospective study of 27 steroid-naive, ambulant male children/adolescents with genetically-confirmed DMD (mean age, 8.8 ± 3.3 years). Functional performance was assessed using motor function measure (MFM) which assess standing/transfer (D1), proximal (D2) and distal (D3) motor function. And six-minute-walk test (6MWT). Imaging evaluation included quantitative muscle MRI which measured muscle fat content in a specific location of right rectus femoris by mDixon sequence. Quantitative MUS measured muscle brightness in standardized US image as an indicator of muscle fat content.

**Results:** We found a highly significant positive correlation between the mean MFM total score and 6MWT (R=0.537, P=0.007). And a highly significant negative correlation between fat content by MUS and MFM total score (R=-0.603, P=0.006) and its D1 subscore (R=-0.712, P=0.001). And a significant negative correlation between fat content by US and 6MWT (R=-0.529, P=0.02). And a significant positive correlation between muscle fat content by mDixon MRI and patient’s age (R=0.617, P=0.01).

**Discussion:** Quantitative MUS correlates significantly with clinical/functional assessment tools as MFM and 6MWT, and augments their role in disease-tracking of DMD. Quantitative MUS has the potential to act as a substitute to functional assessment tools. The role for quantitative muscle MRI in disease-tracking should be further explored after elimination of confounding factors.

**Graphical abstract:** Divergent arrows represent negative correlations, while convergent the arrow represents a positive correlation.

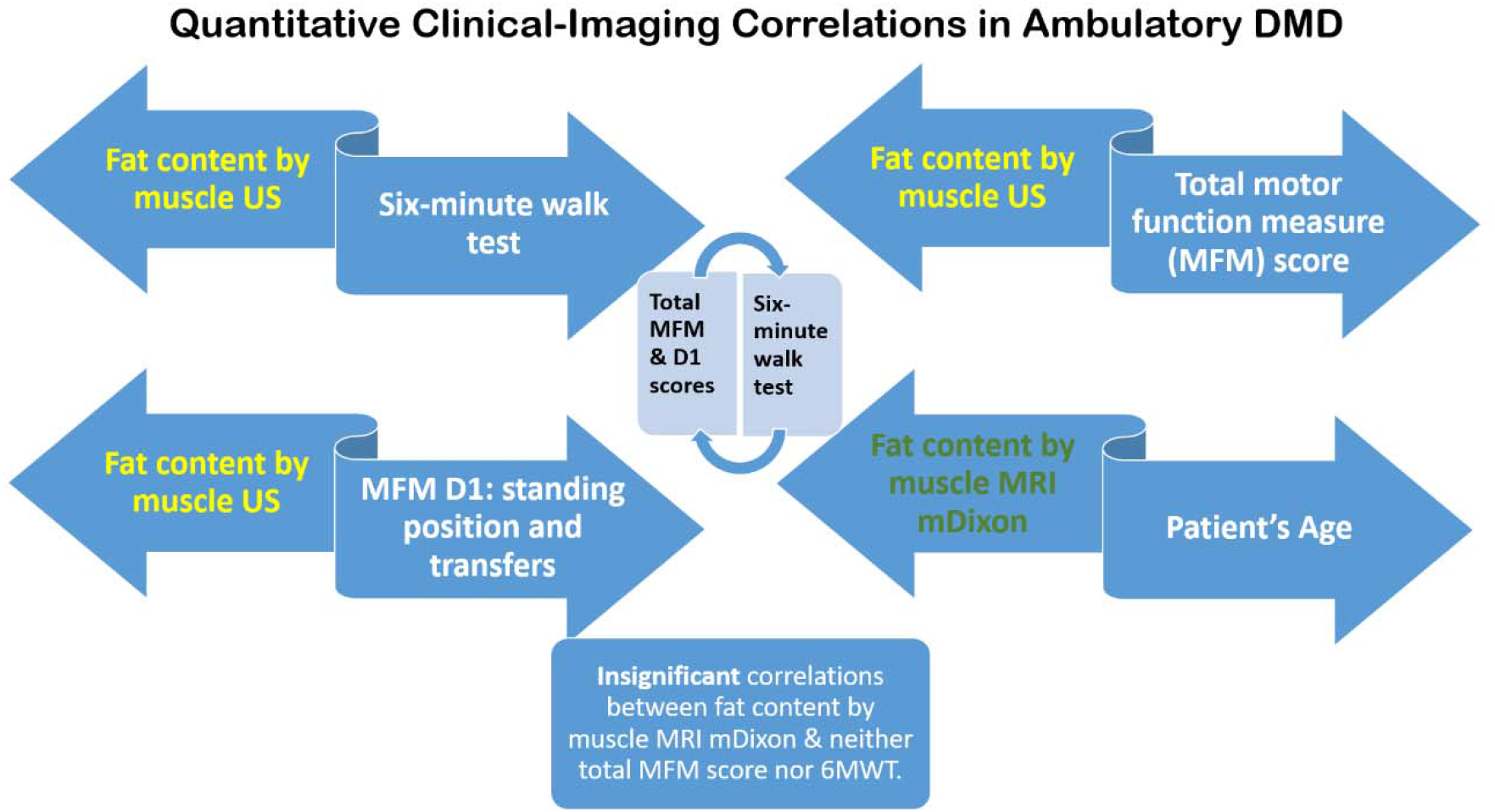

## Introduction

Duchenne muscular dystrophy (DMD) is a severe, progressive X-linked inherited disease that affects 1 in 3600– 6000 live male births (Falzarano et al., 2015). DMD occurs as a result of mutations (mainly deletions) in the dystrophin gene (DMD; locus Xp21.2). The most common presenting feature is secondary deterioration of the motor milestones typically recognized at mid childhood. With time, untreated progressive muscle weakness, joint contractures, and cardio-pulmonary compromise affect quality of life. The disease eventually runs a fatal course by approximately 20 years of age (Suthar and Sankhyan, 2018; Falzarano et al., 2015). The management of DMD requires a multidisciplinary approach to improve the quality of life (Suthar and Sankhyan, 2018; Apkon et al., 2018; Salera et al., 2017).

Therapeutic interventions have recently demonstrated intense upswings in modifying the natural history of DMD. These interventions include; therapies that slow the decline in muscle strength and function as glucocorticoids and therapies that target the pathology of DMD or improve muscle growth and regeneration (Falzarano et al., 2015; Liew and Kang, 2013). Furthermore, gene therapies as gene-addition, exon-skipping, stop codon read through and genome-editing therapies aim at improving the expression and functionality of dystrophin protein. Other therapies work on a cellular level and aim at replacement of damaged muscle tissue. Both of the above therapies yielded encouraging preliminary results (Verhaart et al., 2019; Barthélémy et al., 2018). Nevertheless these therapeutic gains have accentuated the need for consistent, valid and reliable assessment tools/outcome measures to monitor responses to treatment and to act as prognostic indicators. Widely used clinical scales that measure the motor function and activities of ambulatory and non-ambulatory patients with DMD exist (Mcdonald et al., 2013; Mayhew et al., 2011; Bushby et al., 2010). Recently significant interest in the use of magnetic resonance imaging (MRI) of muscles as whole-body MRI, quantitative MRI and magnetic resonance spectroscopy (MRS) to monitor disease progression has evolved (Warman Chardon et al., 2019; Willcocks et al., 2017; Senesac et al., 2015). MRI is a valuable non-invasive tool to reveal distinct pathologic patterns of various hereditary muscular diseases (Aivazoglou et al., 2021) such as facioscapulohumeral muscular dystrophy (Leung, 2017; Fatehi et al., 2017; Gerevini et al., 2016; Leung et al., 2015; Díaz-Manera et al., 2015), sarcoglycanopathies (Tasca et al., 2018), congenital myopathies (Leung 2017; Klein et al., 2011), muscular dystrophies (Sakr et al., 2021; Fu et al., 2016; Díaz-Manera et al., 2015), including DMD (Leung 2017; Polavarapu K et al., 2016; Li W et al., 2015). The use of quantitative muscle MRI and whole-body MRI has also been shown valuable to draw clinical-imaging-genetic correlations and identify ideal sites for muscle biopsy (Warman Chardon et al., 2019; Sakr et al., 2021; Leung 2017; Fatehi et al., 2017; Polavarapu et al., 2016; Gerevini et al., 2016; Li et al., 2015). The combined use of quantitative muscle US and MRI showed promising results in regard to delineating disease stage and its relevance to functional status in facioscapulohumeral muscular dystrophy patients (Mul et al., 2018; Janssen et al., 2014). In DMD quantitative muscle MRI (Nagy et al., 2020; Bonati et al., 2015) and quantitative muscle ultrasound (US) (Zaidman et al., 2017) were found to be a satisfactory substitute to clinical/functional assessment tools as timed function tests in regard to monitoring disease progression.

Nevertheless, we are not aware of studies that investigated the clinical utility of quantitative muscle US and quantitative muscle MRI when used simultaneously with clinical functional tests in patients with DMD. The objectives of this study are: a) to assess the role of quantitative muscle US and muscle MRI as a potential diagnostic and prognostic tool in a series of ambulatory children with genetically-confirmed DMD, b) to draw relevant correlations between these two imaging modalities and clinical outcomes namely 6-minute walk test and motor function measure.

## Material and Methods

This was a prognostic cross-sectional retrospective study. Twenty seven steroid-naive, ambulant male children/adolescents with genetically-confirmed DMD were enrolled. The mean age of patients was 8.8 ± 3.3 years (range, 3.1 to 18). The functional performance of patients was assessed using the motor function measure (MFM) (Vuillerot et al. 2010) and six-minute walk test (6MWT) (Mcdonald et al., 2013). MFM consists of 32 items (20 items for children <7 years). It assesses all three dimensions of motor performance including standing and transfer (D1) subscore, axial and proximal motor function (D2), and distal motor function (D3). The 6MWT a commonly used timed functional test that also sufficiently monitors changes in muscle function. The imaging evaluation included quantitative muscle MRI which measured muscle fat content in the right rectus femoris at the junction of the proximal 1/3 & distal 2/3 of thigh by mDixon sequence and quantitative muscle US which measured muscle brightness in standardized US image as an indicator of muscle fat content. Twenty five patients completed both functional tests, 21 patients completed the muscle US examination and 16 completed the muscle MRI examination. The study was approved by the Medical Ethics Research Committee of Faculty of Medicine, Ain Shams University, Egypt, number FMASU R 110 / 2021. Informed consent from participants or waived by the regulatory authority.

### Assessment tools

The imaging and functional evaluation took place over two consecutive days where imaging evaluations were done prior to the functional tests. Patients who had sufficient cognitive ability to comply with verbal commands pertaining to the functional tests, were evaluated by the same examiner and approximately at the same time-point of the day. The functional tests were performed uniformly in the following order; MFM followed by 6MWT.

#### Motor function measure (MFM)

Items of the MFM-32 and MFM-20 are classified in 3 domains: D1: Standing and transfers (13 items for the MFM-32 and 8 items for the MFM-20); D2: Axial and proximal motor function (12 items for the MFM-32 and 8 items for the MFM-20); D3: Distal motor function (7 items for the MFM-32 and 4 items for the MFM-20). Scores ranged from 0 to 3, as follows: (0) cannot perform the task, or cannot maintain the starting position, (1) initiated the task, (2) performs the movement incompletely, or completely but imperfectly (compensatory movements, position maintained for an insufficient duration of time, slowness, uncontrolled movement) and (3) performs the task fully and “normally” namely the movement is controlled, mastered, directed and performed at constant speed. The calculation of scores was expressed as a percentage in relation to the maximum score https://mfm-nmd.org/.

#### Six-minute walk test

The 6MWT was performed according to the ATS guidelines (ATS Committee, 2002), modified by having two examiners, one recording time and distances and one staying close to the patient for safety issues.

#### Quantitative muscle MRI

mDixon sequence was taken using Achieva 1.5-T MR machine (Philips Medical Systems, The Netherlands) with the following parameters; (Slice thickness 10 mm, Spacing 5 mm, Number of phase encoding steps: 268, Acquisition matrix 272/0/0/268, Flip angle 15), the images were transferred to workstation (ViewForum R 6.3). The mean fat and water signal of a region of interest (ROI) within the right rectus femoris muscle was measured at the junction between the proximal 1/3 and distal 2/3 between its origin (from the anterior inferior iliac spine) and its insertion (at the upper pole of the patella) was measured.

#### Quantitative muscle US

US in axial plane of the right rectus femoris muscle was taken at the junction between the proximal 1/3 and distal 2/3 between its origin (from the anterior inferior iliac spine) and its insertion (at the upper pole of the patella, using GE Logiq p7 machine (GE Healthcare, Waukesha, Wisconsin, USA) with high resolution linear probe 7-12 MHz. All the imaging parameters (including the probe frequency, depth, and gain) were constant, the images were transferred to a personal PC and mean grayscale (i.e. muscle echogenicity) was calculated using image histogram analysis software (ImageJ) and was used as an indication of muscle fat content.

### Statistical methods

Data was revised for its completeness and consistency. Data entry was done on Microsoft Excel workbook. Quantitative data was summarized by mean, standard deviation while qualitative data was summarized by frequencies and percentages. The program used for data analysis was IBM SPSS statistics for windows version 23 (IBM Corp., Armonk, NY, USA). Chi-square test, student t test, and Pearson correlation coefficient were used in analysis of this study. Kappa test was done to measure level of agreement. A “P value” of less than 0.05 was considered statistically significant.

## Results

Six patients 22% were < 7 years of age. 19 patients (70%) had exon deletions, 2 (7%) had exon duplications, and 6 (22%) had small mutations. Comparatively, patients with exon deletions had a lower mean MFM score without statistical significance. Tables 1 and 2 show the descriptive statistics of all clinical and imaging assessment tools used. We found a highly significant positive correlation between the 6MWT and the mean total MFM score (R=0.537, P=0.007) and its D1 subscore (R=0.751, P=0.000) (Figs. 1, 2). We found a highly significant negative correlation between fat content by muscle US and total MFM score (R=-0.603, P=0.006) and its D1 subscore (R=-0.712, P=0.001) (Figs. 3, 4, 5). Additionally, we found a significant negative correlation between fat content by muscle US and 6MWT score (R=-0.529, P=0.02) (Table 3). This denotes that the higher the fat replacement in muscles of the thigh, the lower the scores of MFM and its D1 subscore and the lower meters achieved by patients in 6MWT. We found a significant positive correlation between patient’s age and muscle fat content by mDixon MRI (R=0.617, P=0.01). However, we did not find a statistically significant correlation between both muscle fat content and muscle water content by mDixon MRI and neither total MFM score nor 6MWT (Table 4) (Figs. 6, 7) (Fig. S8). Statistical correlations between age and both mean total MFM scores and subscores, and 6MWT scores were non-significant. For additional information see (File S1).

**Table 1.** Descriptive statistics of functional assessment tools.

|  | N | Mean | SD | IQR | Median | Min | Max |
| --- | --- | --- | --- | --- | --- | --- | --- |
| Age | 27 | 8.7 | 3.4 | 6.6-10.2 | 8.6 | 3.1 | 18 |
| **MFM** |  |  |  |  |  |  |  |
| Total% | 25 | 74.3 | 17.6 | 63.9-88.0 | 78.3 | 26.0 | 96.8 |
| D1% | 25 | 57.7 | 26.8 | 37.1-76.9 | 61.5 | 0 | 97.4 |
| D2% | 25 | 86.2 | 17.3 | 83.3-97.2 | 94.4 | 30.5 | 100 |
| D3% | 25 | 84.8 | 13.5 | 75.5-95.2 | 85.7 | 42.8 | 100 |
| **6MWT**<br>(meters) | 25 | 291.4 | 112.4 | 241.5-370 | 304.0 | 40 | 525 |

N: Number of patients, MFM: motor function measure, 6MWT: 6-minute walk test, SD: Standard Deviation, IQR: Interquartile range, Min: Minimum, Max: Maximum

**Table 2.** Descriptive statistics of imaging assessment tools.

|  | N | Mean | SD | IQR | Median | Min | Max |
| --- | --- | --- | --- | --- | --- | --- | --- |
| US | 21 | 83.5 | 29.1 | 57.0-108.6 | 84.2 | 43.6 | 141.4 |
| mDixon fat | 16 | 77.9 | 60.7 | 44.1-86.5 | 61.6 | 30.2 | 272 |
| mDixon water | 16 | 375.1 | 145.7 | 321.2-468.7 | 431.3 | 52.1 | 535.7 |

N: Number of patients, SD: Standard Deviation, IQR: Interquartile range, Min: Minimum, Max: Maximum

**Table 3.**
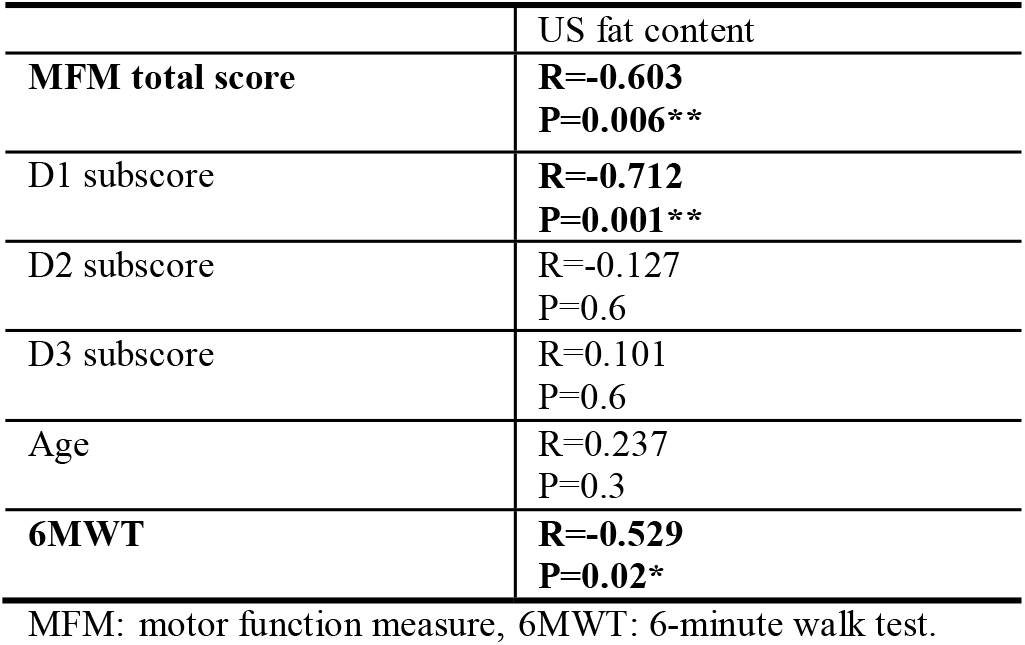
Correlation coefficient between age, functional assessment tools namely MFM and 6MWT and muscle fat content by ultrasound.

|  | US fat content |
| --- | --- |
| **MFM total score** | **R=-0.603**<br>**P=0.006\*\*** |
| D1 subscore | **R=-0.712**<br>**P=0.001\*\*** |
| D2 subscore | R=-0.127<br>P=0.6 |
| D3 subscore | R=0.101<br>P=0.6 |
| Age | R=0.237<br>P=0.3 |
| **6MWT** | **R=-0.529**<br>**P=0.02\*** |

MFM: motor function measure, 6MWT: 6-minute walk test.

**Table 4.**
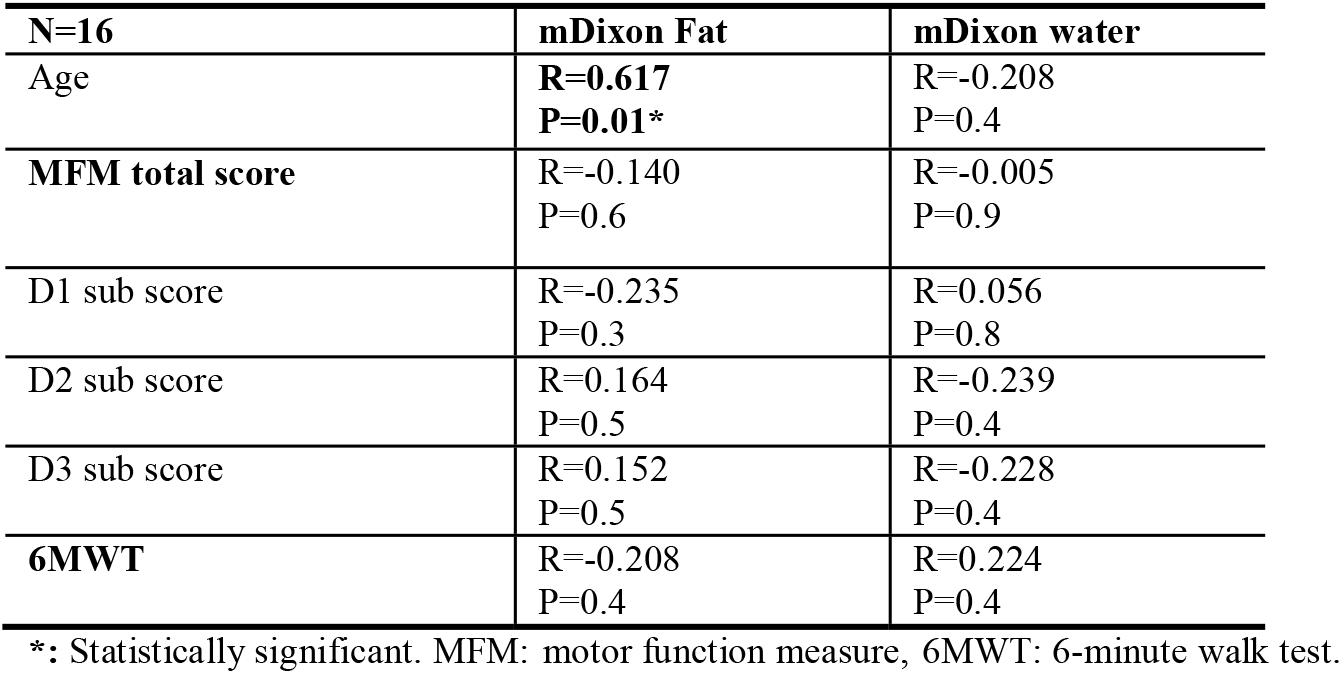
Correlation coefficient between age, functional assessment tools namely MFM and 6MWT and muscle fat and water content by mDixon MRI.

**Figure 1:**
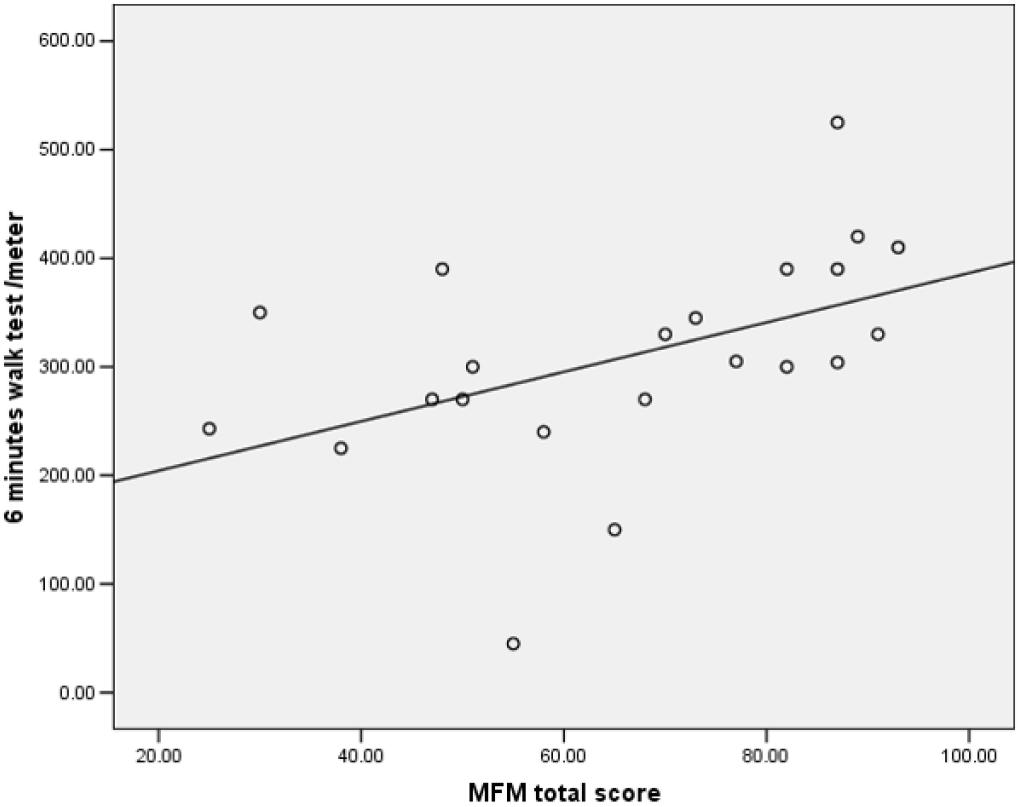
Correlation coefficient between Motor Function Measure (MFM) total score and 6-minute walk test showed a -P<0.01- a highly significant **positive** correlation.

**Figure 2:**
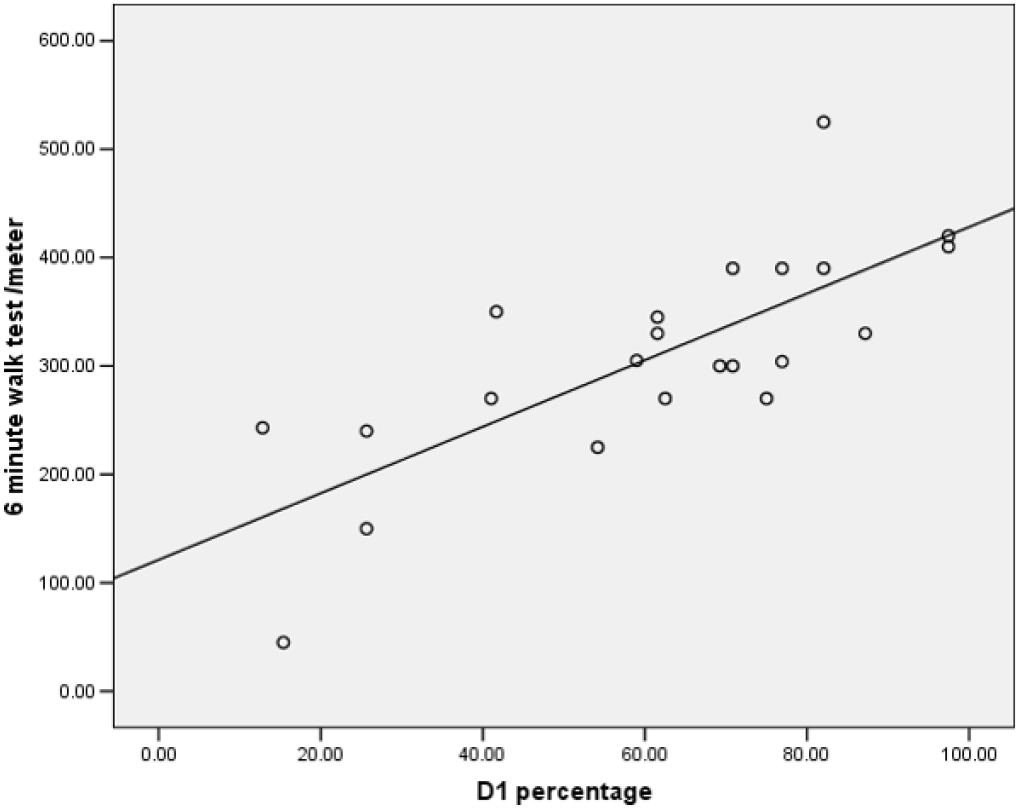
Correlation coefficient between motor function measure (MFM) D1 subscore percentage and 6-minute walk test showed -P<0.01- a highly significant **positive** correlation.

**Figure 3:**
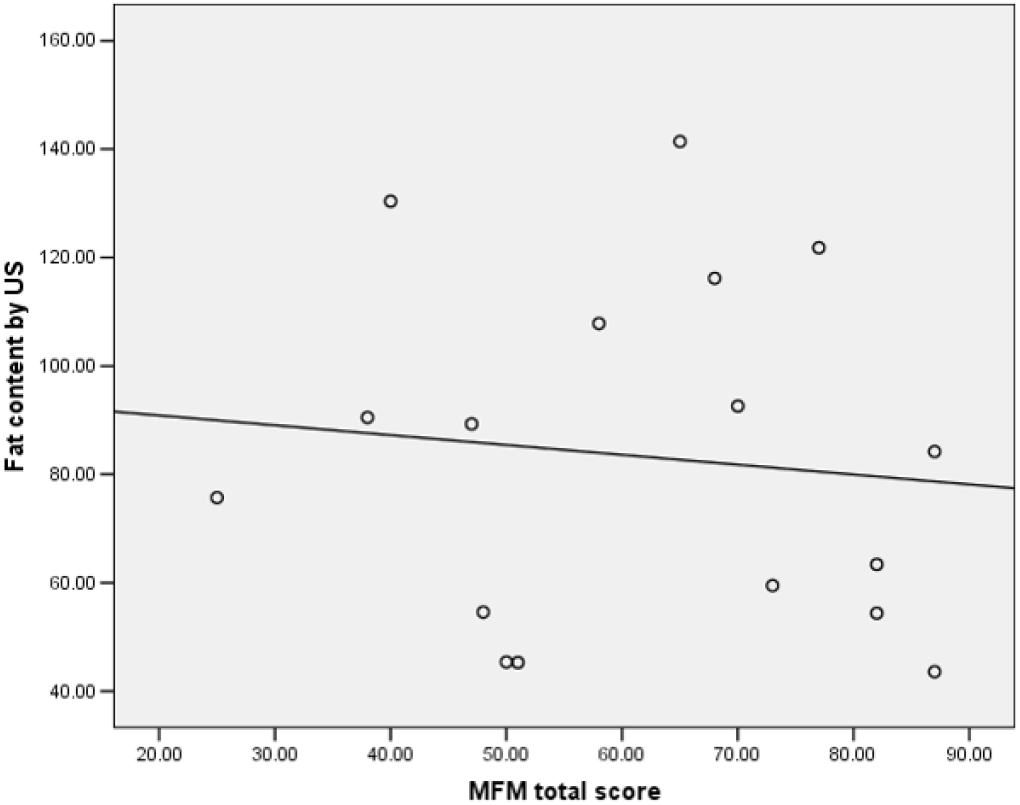
Correlation coefficient between motor function measure (MFM) total score and fat content by muscle ultrasound examination Showed a -P<0.05- a significant **negative** correlation.

**Figure 4:**
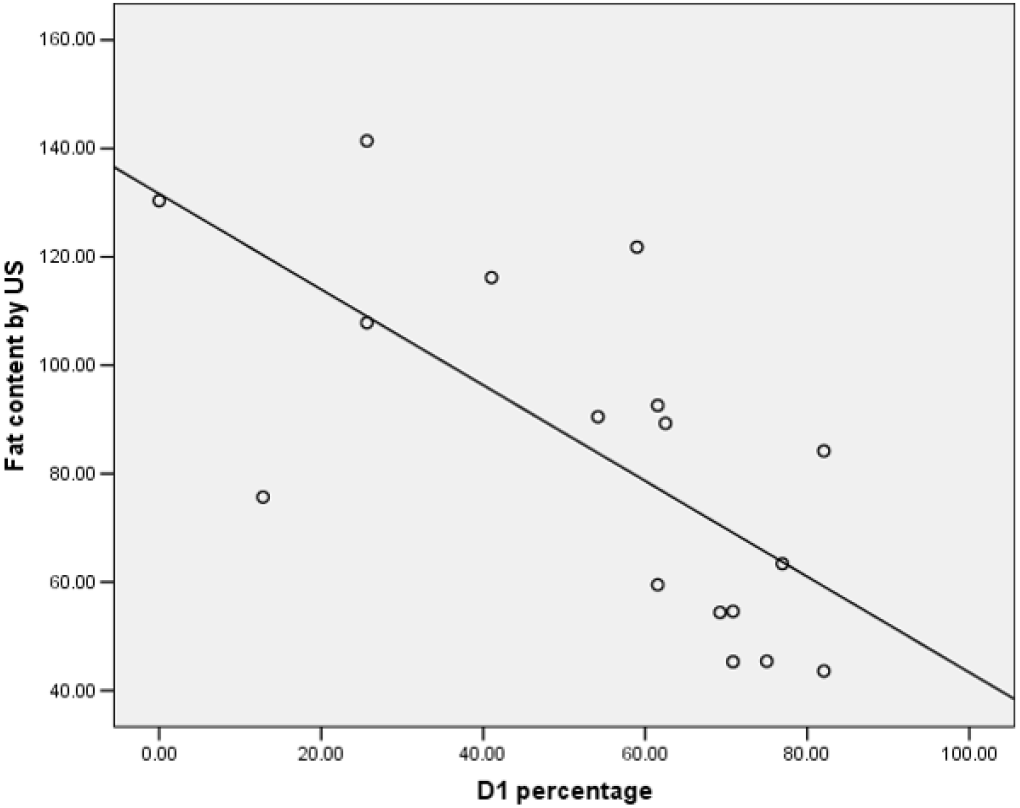
Correlation coefficient between motor function measure (MFM) D1 subscore percentage and fat content by muscle ultrasound examination -P<0.01- a highly significant **negative** correlation.

**Figure 5. (A-C):**
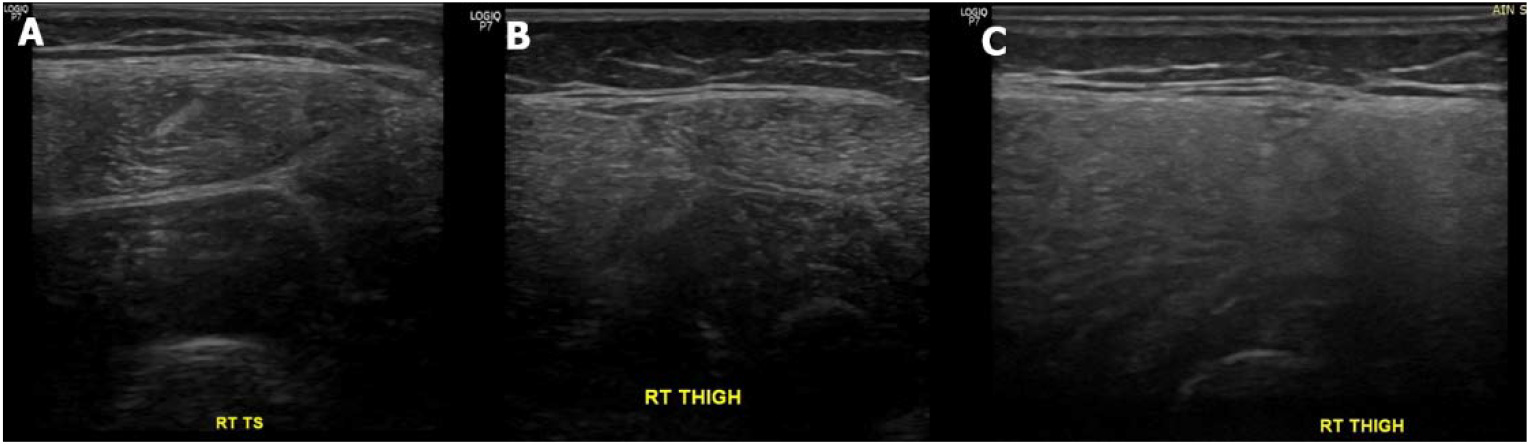
Transverse US image of the right thigh of three different patients (A, B & C) showing different degrees of fatty infiltration of the quadriceps muscle as per increased echogenicity and blurred cortical surface of the femur. The mean histogram is 53, 76 & 80 respectively.

**Figure 6. (A-D):**
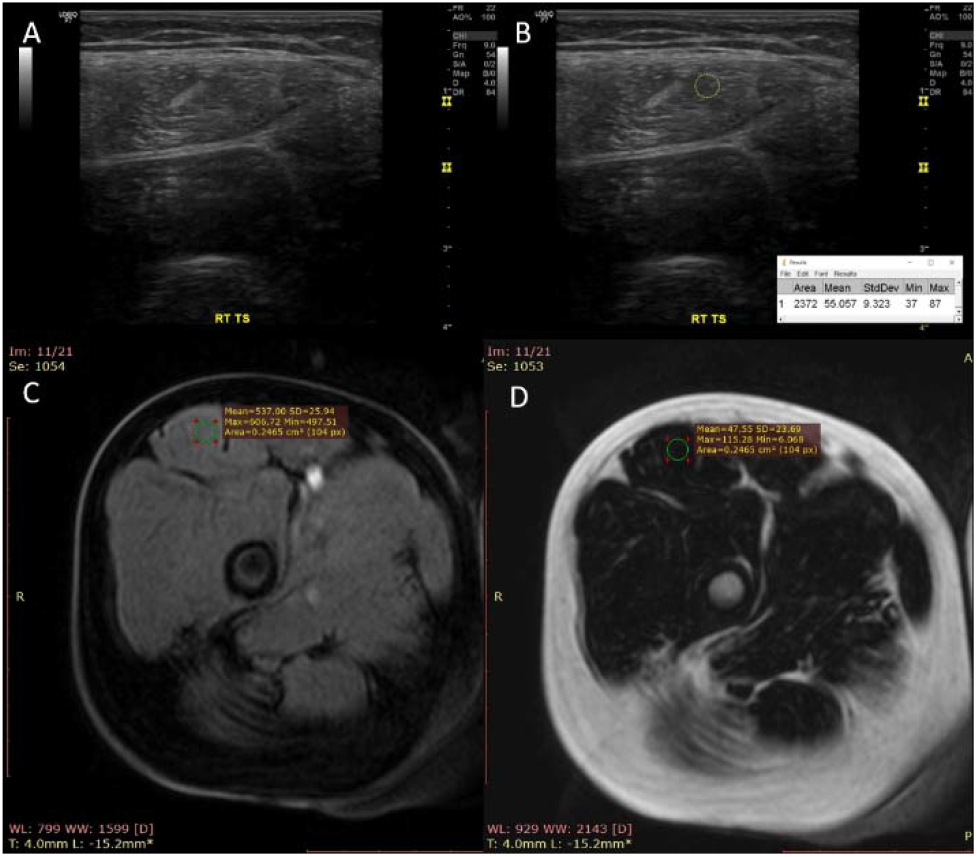
A patient with Duchenne muscular dystrophy. (A) Transverse US image of the right thigh taken at the junction between the proximal 1/3 and distal 2/3 between its origin (from the anterior inferior iliac spine) and its insertion (at the upper pole of the patella) showing fatty infiltration of the quadriceps muscle as shown by increased echogenicity and blurred cortical surface of the femur. (B) The same image in (A) analyzed by ImageJ software with the ROI is within the rectus femoris muscle, mean histogram, 55. (C, D) Axial MRI image (mDixon sequence) with the region of interest (ROI) within the right rectus femoris muscle at the junction between the proximal 1/3 and distal 2/3 between its origin (from the anterior inferior iliac spine) and its insertion (at the upper pole of the patella), mean fat & water signals were 66 & 546 respectively.

**Figure 7. (A-F):**
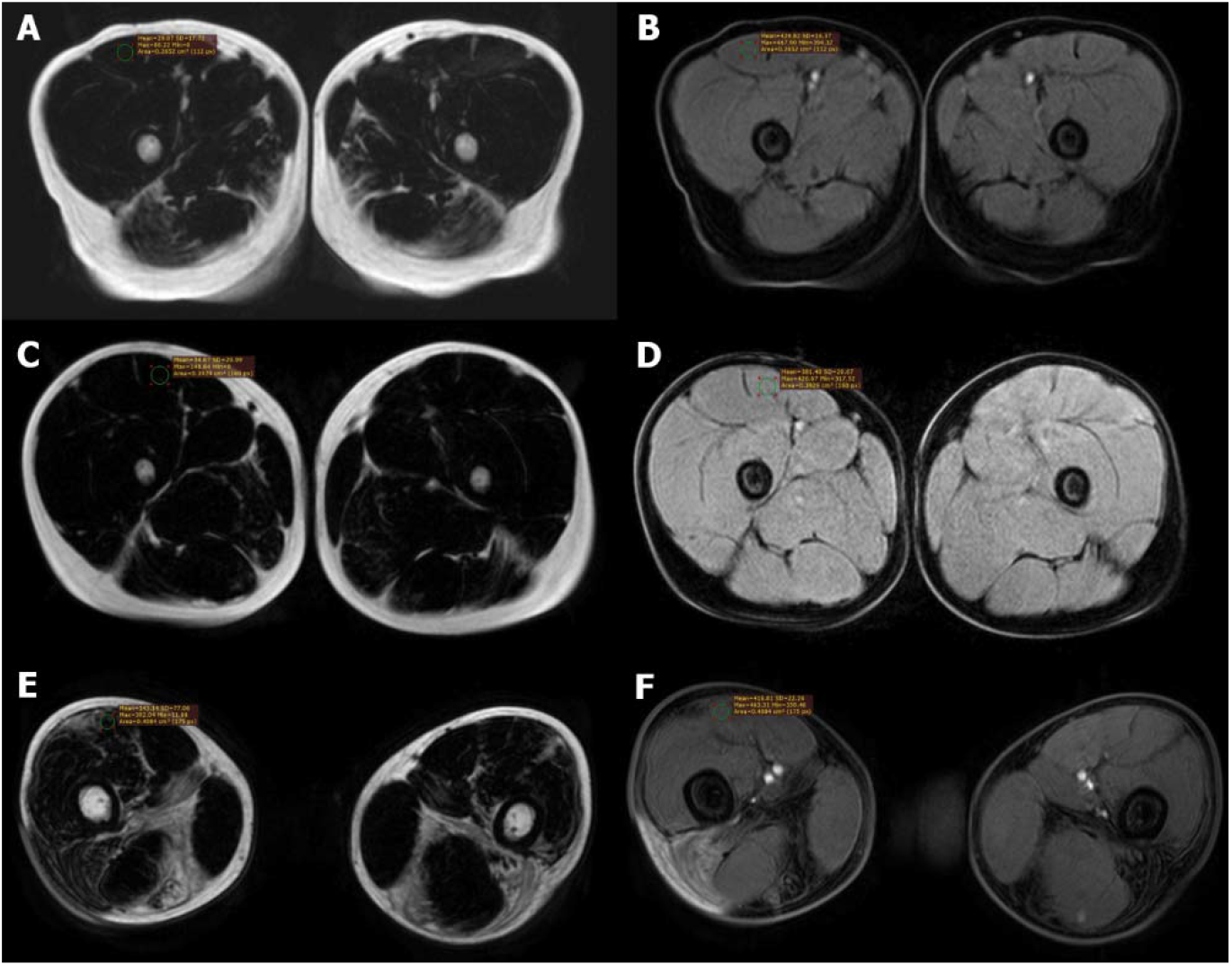
Axial MRI image (mDixon sequence) in three different patients with DMD showing fat (A, C & E) and water (B, D & F) signal. The mean fat signals were 29, 35 & 143 respectively while those of water were 430, 351 & 416 respectively.

## Discussion

Corticosteroids remain the mainstay of treatment in DMD children whereas gene therapeutic modalities are emerging (Rivera et al., 2020; Frank et al., 2020) among others (Nagy et al., 2019). Both treatment modalities aim at improving the child’s functional status. Recent advances in therapeutics for the treatment of DMD children have sparked significant interest in finding reliable clinical and imaging assessment tools to monitor responsiveness to these treatment modalities. The current study has incorporated various clinical and imaging assessment tools to explore their diagnostic and prognostic role in genetically-confirmed ambulatory and steroid-naïve DMD children and adolescents. Whereas quantitative US depends on measuring muscle echogenicity using the mean gray scale of a ROI within a selected muscle (Zaidman et al., 2017), quantitative MRI depends on measuring muscle fat replacement (Bonati et al., 2015; Burakiewicz et al., 2017). Both quantitative imaging tools have proved to aid clinical assessment fundamentally.

Our study implications are twofold. Firstly, it confirmed the widely acknowledged role of MFM and 6MWT as reliable and clinically meaningful assessment tools in DMD children (Nagy et al., 2020; Vuillerot et al., 2010; Mcdonald et al., 2013). Secondly, our study introduced a statistically significant positive correlations between scores of these clinical assessments tools namely MFM and 6MWT on the one hand and quantitative muscle US in DMD children. This highlights the potential clinical utility of quantitative muscle US for DMD monitoring. And underscores the role of quantitative muscle US as an important substitute and supplement to clinical functional assessment tools. This role has been elucidated in both cross-sectional (Shklyar et al., 2015) and longitudinal (Zaidman et al., 2017) study designs capable of monitoring disease progression in DMD children. Contrastingly, quantitative muscle fat content measured by US did not show a statistically significant correlation with patient’s age.

Quantitative muscle MRI -as per T2 mapping and mDixon- has been found helpful for delineating upper (Forbes et al., 2020; Willcocks et al., 2017) and lower extremity (Naarding et al., 2020; Yin et al., 2019; Bonati et al., 2015) muscle involvement and to be a satisfactory predictor of both strength of the investigated muscles or of the overall function of the investigated extremity in DMD children. However, our results did not establish a similarly significant correlation between the scores of these clinical assessments tools namely MFM and 6MWT and quantitative muscle MRI namely muscle fat content and muscle water content by mDixon MRI. This may be attributed to the limitations of our study. Our study contained multiple clinical and imaging assessment tools (dependent variables). Additionally, its retrospective and cross-sectional nature does not allow for complete bias control in terms of standardization of patient characteristics among others. Moreover, longitudinal study designs have a greater potential to consolidate evidence for the use of both quantitative muscle US and muscle MRI as a substitute or a supplement to functional assessment tools in disease tracking and monitoring response to treatment of DMD patients. Further, the diversity of clinical assessment tools in general and functional tools in specific used across studies needs to be considered when interpreting the imaging-clinical correlation results (Senesac et al., 2019). Interestingly, quantitative muscle fat content measured by mDixon MRI showed a statistically positive correlation with patient’s age. This is a clinically meaningful correlation. And it supports the assumption that the above-noted non-significant correlations between the clinical MFM and 6MWT scores, and quantitative fat content by mDixon MRI is mainly due study limitaions. Additionally, of note is that statistical significance does not necessary equate to clinical significance. Consequently all findings should be interpreted cautiously and within clinical context.

## Conclusions

Quantitative muscle US correlates significantly with clinical/functional assessment tools as MFM and 6MWT, and augments their promising role in disease tracking of DMD children. In that regard quantitative muscle US has the potential to act as substitute or supplement to functional assessment tools. The potential role for all components of quantitative muscle MRI in disease tracking should be further explored after elimination of confounding factors. The presence of multiple clinical and imaging assessment tools (dependent variables) and study design-related limitations may have underpowered our statistical correlations.

## Data Availability

The authors confirm that most of the data supporting the findings of this study are available within the article and its supplementary materials.
Additional data that support the findings of this study are available on request from the corresponding author, [HMS]. The data are not publicly available due to [restrictions e.g. their containing information that could compromise the privacy of research participants].

## Acknowledgements

The authors thank Dr Khaled M. Abd Elaziz, Professor of Public Health, Ain Shams University for his help in performing the statistical analysis.

## Supplementary Information

**Figure S8:** Transverse US image of the right thigh of three different children with DMD

**File S1: Tables S5 to S25**

