## Supplementary material for "Ambulatory Duchenne Muscular Dystrophy Children: Cross-sectional Correlation between Function, Quantitative Muscle Ultrasound and MRI": (File S1)

**Hala Abdulhady MD^1^,**
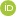
**<https://orcid.org/0000-0002-3987-0658> Hossam M. Sakr MD^2*^**
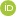
**<https://orcid.org/0000-0002-7055-5736> Nermine S. Elsayed MD^3^,**
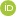
**<https://orcid.org/0000-0001-6510-9419> Tamer A. El-Sobky MD^4^,**
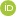
**<https://orcid.org/0000-0001-8670-0419> Nagia Fahmy MD^5^,**
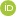
**<https://orcid.org/0000-0002-8964-4923> Amr M. Saadawy MD^2^, Heba Elsedfy MD^3^**

^1^Department of Physical Medicine, Rheumatology and Rehabilitation, ^2^Department of Diagnostic and Interventional Radiology and Molecular Imaging, ^3^Department of Medical Genetics, ^4^Division of Pediatric Orthopedics, Department of Orthopedic Surgery, ^5^Neuromuscular Unit, Department of Neuropsychiatry, **Faculty of Medicine, Ain Shams University, Cairo, Egypt**

**Motor function measure (MFM) and 6-minute walk test correlations**

**Tables: S5 to S25**

**Table S5:** Age distribution of studied patients

| N=27 | No. | % |  |
| --- | --- | --- | --- |
| **Age**  Below 7 years  7 years or more | 7  20 | 25.9  **74.1** |  |
|  | Mean | SD | Range |
| **Age (years)** | 8.7 | 3.4 | 3.1-18 |
| **Median age** | **8.6** |  |  |

This table shows that 74.1% of the patients were 7 years or more of age. The mean age of patients was 8.7 years while the median age is 8.6 years.

**Table S6:** Distribution of type of mutation by molecular findings

| N=27 | No. | % |
| --- | --- | --- |
| Deletion | 19 | **70.4** |
| Duplication | 2 | 7.4 |
| **Small mutation** | 6 | 22.2 |

This table shows that 70.4% of the cases had gene deletion and 22.2% had small mutation while 7.4% of the patients had duplication.

**Table S7: Comparison between gene abnormality and the mean age of patients**

| Age | Mean | SD | F | P |
| --- | --- | --- | --- | --- |
| Deletion N=19 | **9.3** | 3.9 |  |  |
| Duplication N=2 | 7.5 | 0.07 | 0.7 | 0.4 |
| **Small mutation N=6** | 7.6 | 1.4 |  |  |

P>0.05 not significant. This table shows a higher mean age among patients with deletion compared to other groups with no significant difference statistically. This table shows a lower mean age among cases with duplication or small mutation compared to deletion group with no statistically significant difference.

**Table S8:** Comparison between age of patients and type of gene mutation

| **Age** | Deletion  No. % | Duplication  No. % | Small mutation  No. % | X2 | P |
| --- | --- | --- | --- | --- | --- |
| Below 7 N=7 | 5 71.4 | 0 | 1 28.6 | 0.8 | 0.6 |
| 7 or more N=20 | 14 70.0 | 2 **10.0** | 4 20.0 |  |  |

P>0.05 not significant. This table shows a higher percentage of small mutation among cases in group of 7 or above compared to cases below 7 years of age but the difference is not significant statistically.

**Table S9:** Comparison between the age groups in regard to the mean MFM D1 subscore.

| **D1 percentage** | Mean | SD | t | P |
| --- | --- | --- | --- | --- |
| Below 7 N=7 | 62.5 | 12.5 | 0.5 | 0.8 |
| 7 or more N=20 | 55.8 | 30.7 |  |  |

P>0.05 not significant. MFM: motor function measure. This table shows no statistically significant difference between the two age groups as regards D1 subscore of the motor function measure. A higher mean D1 subscore in younger age group compared to older age group with no statistically significant difference.

**Table S10:** Comparison between the age groups in regard to the mean MFM D2 subscore.

| **D2 percentage** | Mean | SD | t | P |
| --- | --- | --- | --- | --- |
| Below 7 N=7 | 81.5 | 17.1 | 0.8 | 0.4 |
| 7 or more N=20 | 88.1 | 17.5 |  |  |

P>0.05 not significant. MFM: motor function measure. This table shows a higher mean D2 subscore among older age group with no statistically significant difference.

**Table S11:** Comparison between the age groups in regard to the mean MFM D3 subscore.

| **D3 percentage** | Mean | SD | t | P |
| --- | --- | --- | --- | --- |
| Below 7 N=7 | 79.7 | 9.4 | 1.1 | 0.2 |
| 7 or more N=20 | 86.7 | 14.5 |  |  |

P>0.05 not significant. MFM: motor function measure. This table shows a higher mean D3 sub score among older age group patients with no statistically significant difference.

**Table S12**: comparison between the age groups in regard to the mean MFM total score.

| **MFM total score (%)** | Mean | SD | t | P |
| --- | --- | --- | --- | --- |
| Below 7 N=7 | 73.5 | 13.2 | 0.1 | 0.8 |
| 7 or more N=20 | 74.7 | 19.3 |  |  |

P>0.05 not significant. MFM: motor function measure. This table shows no significant difference statistically between the two age groups as regards the mean total motor function measure percentage.

**Table S13:** Comparison between the age groups as regards the 6-minute walk test (6MWT)

| **6MWT** | Mean | SD |  | t | P |
| --- | --- | --- | --- | --- | --- |
| Below 7 N=7 | 302.0 | 54.8 |  | 0.2 | 0.7 |
| 7 or more N=18 | 287.3 | 129.3 |  |  |  |

P>0.05 not significant. This table shows no significant difference statistically between the two age groups as regards the mean meters of the 6-minute walk test. The table shows a lower mean value for 6-minute walk test among cases above seven years of age compared to younger age group with no statistically significant difference.

**Table S14: Correlation coefficient between the age and MFM scores i.e. total score and subscores.**

|  | **Age** |
| --- | --- |
| **MFM total score** | R=0.044  P=0.8 |
| D1 subscore | R=-0.007  P=0.9 |
| D2 subscore | R=0.063  P=0.7 |
| D3 subscore | R=0.163  P=0.4 |

P>0.05 not significant. This table shows no significant correlation between the total MFM score or the subscores and the age of DMD patients.

**Table S15:** Distribution of rapid decline of ambulation according to the 6-minute walk test. Cut off value 350 meters.

| **N=25** | No. % | 95% CI |
| --- | --- | --- |
| No Rapid decline | **7 28** | **12.0-49.0** |
| Rapid decline | **18 72** | **50.6-87.9** |

This table shows that 72% of the patients have rapid decline in ambulation in the next 48 months as predicted by the results of the 6-minute walk test below 350 m while 28% have no rapid decline in ambulation.

**Table S16:** Distribution of rapid decline of ambulation according to the 6-minute walk test (Cut off value 300 meters)

| **N=25** | No. % | 95% CI |
| --- | --- | --- |
| No Rapid decline | **15 60** | **38-78** |
| Rapid decline | **10 40** | **21-61** |

This table shows that 40% of the patients have rapid decline in ambulation in the next 48 months as predicted by the results of the 6-minute walk test below 300 m while 60% have no rapid decline in ambulation.

**Table S17: Correlation coefficient between the MFM results, age, 6-minute walk test and mDixon fat and water findings of MRI**

| **N=16** | **mDixon Fat** | **mDixon water** |
| --- | --- | --- |
| Age | **R=0.617**  **P=0.01*** | R=-0.208  P=0.4 |
| **MFM total score** | R=-0.140  P=0.6 | R=-0.005  P=0.9 |
| D1 sub score | R=-0.235  P=0.3 | R=0.056  P=0.8 |
| D2 sub score | R=0.164  P=0.5 | R=-0.239  P=0.4 |
| D3 sub score | R=0.152  P=0.5 | R=-0.228  P=0.4 |
| 6MWT | R=-0.208  P=0.4 | R=0.224  P=0.4 |

*P<0.05 significant. This table shows a significant positive correlation between mDixon fat values and age of patients. This table shows no correlation between mDixon fat values and MFM, 6-minute walk test values. This table shows no correlation between mDixon water content and MFM, 6-minute walk test.

**Table S18:** Correlation coefficient between the MFM results, age, 6-minute walk test and MRS Press and MRS steam findings of MRI

| **N=16** | **MRS Press** | **MRS steam** |
| --- | --- | --- |
| Age | R=0.058  P=0.8 | R=0.047  P=0.8 |
| **MFM total score** | **R=0.651**  **P=0.009**** | **R=0.660**  **P=0.01*** |
| D1 sub score | **R=0.594**  **P=0.03*** | R=0.469  P=0.09 |
| D2 sub score | **R=0.538**  **P=0.03*** | R=0.393  P=0.1 |
| D3 sub score | **R=0.534**  **P=0.04*** | **R=0.791**  **P=0.001*** |
| 6MWT | R=0.373  P=0.1 | R=0.310  P=0.2 |

*P<0.05 Significant, **P<0.01 highly significant.

This table shows a highly significant positive correlation between MRS press values and total MFM score.

This table shows a significant positive correlation between MRS press values and D1, D2, D3 subscores of MFM. This table shows a significant positive correlation between MRS steam and total MFM score. This table shows a highly **significant positive correlation** between D3 subscore and the MRS steam findings of MRI

**Table S19:** Correlation coefficient between the MFM results, age, 6-minute walk test and DTI finding of MRI results

| **N=14** | DTI. FA | DTI. ADC |
| --- | --- | --- |
| **MFM total score** | R=-0.438  P=0.1 | R=0.417  P=0.1 |
| D1 sub score | **R=-0.640**  **P=0.01*** | **R=0.617**  **P=0.01*** |
| D2 sub score | R=-0.283  P=0.3 | R=0.246  P=0.3 |
| D3 sub score | R=0.004  P=0.9 | R=0.032  P=0.9 |
| age | R=0.402  P=0.1 | R=-0.404  P=0.1 |
| 6MWT | **R=-0.701**  **P=0.004**** | **R=0.782**  **P=0.001**** |

*P<0.05 Significant, **P<0.01 highly significant. This table shows a significant negative correlation of D1 subscore and the DTI FA findings. Which denotes the higher D1 scores associated with lower DTI FA scores. This table shows a significant positive correlation of D1 subscore and the DTI ADC findings. Which denotes the higher D1 scores associated with high DTI ADC	scores.

**Table S20:** Comparison between type of gene mutation and the screening of patients with 6-minute walk test. Cut off value 300

| **300** | No rapid decline  No. % | Rapid decline  No. % | X2 | P |
| --- | --- | --- | --- | --- |
| Deletion N=18 | 9 50.0 | 9 **50.0** | 2.9 | 0.08 |
| Others  N=7 | 6 85.7 | 1 14.3 |  |  |

P>0.05 not significant. This table shows a higher percentage of rapid decline among patients with deletion 50% compared to 14.3% among patients with other gene mutation with border line significance.

**Table S21:** Comparison between type of gene mutation and the screening of patients with 6-minute walk test. Cut off value 350

| **350** | No rapid decline  No. % | Rapid decline  No. % | X2 | P |
| --- | --- | --- | --- | --- |
| Deletion N=18 | 4 22.2 | 14 **77.8** | 1.0 | 0.3 |
| Others  N=7 | 3 42.9 | 4 57.1 |  |  |

P>0.05 not significant. This table shows a higher percentage of rapid decline among patients with small mutation or duplication 85.7% compared to 50% among patients with deletion with border line significance.

**Table S22:** Correlation coefficient between the ultrasound findings of fat content and the MRI findings of patients with DMD

|  | US fat content |
| --- | --- |
| **mDixon Fat** | **R=0.743**  **P=0.002**** |
| MRS Steam | R=-0.139  P=0.6 |
| MRS Press | R=0.294  P=0.3 |
| DTI FA | R=0.396  P=0.1 |
| DTI ADC | R=-0.342  P=0.2 |

*P<0.05 Significant, **P<0.01 highly significant. This table shows a highly significant positive correlation between ultrasound fat content and the mDixon fat content among patients with DMD. This table shows no correlation between ultrasound fat content and MRS steam, MRS press, DTI FA and DTI ADC.

**Table S23:** Comparison between type of gene mutation and the ultra sound grouping of patients (fat content of muscles). Cut off value 84 (median value of US)

|  | Low fat content  No. % | High fat content  No. % | X2 | P |
| --- | --- | --- | --- | --- |
| Deletion N=13 | 6 46.2 | 7 53.8 | 0.02 | 0.8 |
| Others  N=8 | 4 50.0 | 4 50.0 |  |  |

P>0.05 not significant

**Table S24:** Comparison between US fat content and the screening of patients with 6-minute walk test. Cut off value 300 for screening test

| **US** | No rapid decline  No. % | Rapid decline  No. % | X2 | P |
| --- | --- | --- | --- | --- |
| Low fat N=10 | 7 70.0 | 3 30.0 | 2.5 | 0.1 |
| High fat  N=9 | 3 33.3 | 6 **66.7** |  |  |

P>0.05 not significant. This table shows a higher percentage of rapid decline among patients with high fat content in US 66.7% compared to 30% among patients with low fat content and the difference is border line significant.

**Table S25:** Comparison between US fat content and the screening of patients with 6-minute walk test. Cut off value 350 for screening test

| **350** | No rapid decline  No. % | Rapid decline  No. % | X2 | P |
| --- | --- | --- | --- | --- |
| Low fat N=10 | 3 30.0 | 7 70.0 | 1.0 | 0.3 |
| High fat  N=9 | 1 11.1 | 8 88.9 |  |  |

P>0.05 not significant. This table shows a higher percentage of rapid decline among patients with high fat content 88.9% compared to 70% among patients with low fat content with no significant difference statistically.
