## Supplementary material for "Ambulatory Duchenne Muscular Dystrophy Children: Cross-sectional Correlation between Function, Quantitative Muscle Ultrasound and MRI": (Fig. S8)

**Hala Abdulhady^1^,**
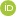
**<https://orcid.org/0000-0002-3987-0658> Hossam M. Sakr^2*^**
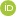
**<https://orcid.org/0000-0002-7055-5736> Nermine S. Elsayed^3^,**
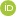
**<https://orcid.org/0000-0001-6510-9419> Tamer A. El-Sobky^4^,**
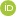
**<https://orcid.org/0000-0001-8670-0419> Nagia Fahmy^5^,**
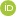
**<https://orcid.org/0000-0002-8964-4923> Amr M. Saadawy^2^, Heba Elsedfy^3^**

^1^Department of Physical Medicine, Rheumatology and Rehabilitation, ^2^Department of Diagnostic and Interventional Radiology and Molecular Imaging, ^3^Department of Medical Genetics, ^4^Division of Pediatric Orthopedics, Department of Orthopedic Surgery, ^5^Neuromuscular Unit, Department of Neuropsychiatry, **Faculty of Medicine, Ain Shams University, Cairo, Egypt**

**^*^**Corresponding:

**Study Highlights**

**1**. Quantitative muscle US correlates significantly with clinical/functional assessment tools as MFM and 6MWT in DMD children.

**2**. Quantitative muscle US has a potential to augment and provide a surrogate for clinical motor functional status in DMD children.

**3**. The potential role for all components of quantitative muscle MRI in disease tracking should be further explored following elimination of confounding factors.

**Figures**

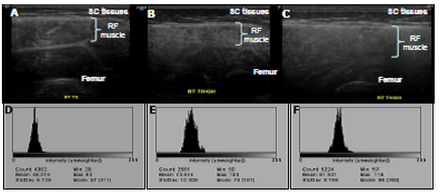

**Figure S8**: Transverse US image of the right thigh of three different children with DMD (A, B & C) showing normal muscle echogenicity grade I Heckmatt score in (A), increased muscle echogenicity grade II Heckmatt score in (B) and increased muscle echogenicity & blurred cortical surface of the femur grade III Heckmatt score in (C), their corresponding histogram (D, E & F) in which the mean is 46, 74 and 82 respectively.
